# The impact of current and future control measures on the spread of COVID-19 in Germany

**DOI:** 10.1101/2020.04.18.20069955

**Authors:** Maria Vittoria Barbarossa, Jan Fuhrmann, Jan H. Meinke, Stefan Krieg, Hridya Vinod Varma, Noemi Castelletti, Thomas Lippert

## Abstract

The novel coronavirus (SARS-CoV-2), identified in China at the end of December 2019 and causing the disease COVID-19, has meanwhile led to outbreaks all over the globe with about 2.2 million confirmed cases and more than 150,000 deaths as of April 17, 2020 [37]. In view of most recent information on testing activity [32], we present here an update of our initial work [4]. In this work, mathematical models have been developed to study the spread of COVID-19 among the population in Germany and to asses the impact of non-pharmaceutical interventions. Systems of differential equations of SEIR type are extended here to account for undetected infections, as well as for stages of infections and age groups. The models are calibrated on data until April 5, data from April 6 to 14 are used for model validation. We simulate different possible strategies for the mitigation of the current outbreak, slowing down the spread of the virus and thus reducing the peak in daily diagnosed cases, the demand for hospitalization or intensive care units admissions, and eventually the number of fatalities. Our results suggest that a partial (and gradual) lifting of introduced control measures could soon be possible if accompanied by further increased testing activity, strict isolation of detected cases and reduced contact to risk groups.

## 1 Introduction

At the end of December 2019, several cases of acute respiratory syndrome were first reported in Wuhan City (Hubei region, China) by Chinese public health authorities. A novel coronavirus was soon found as the main causative agent. This is now known as severe acute respiratory syndrome coronavirus 2 (SARS-CoV-2). The disease caused by SARS-CoV-2, which rapidly spread first through China and then to other countries, is now referred to as coronavirus disease 2019 (COVID-19). The World Health Organization declared COVID-19 a global pandemic on March 11, 2020 [11, 37]. As of April 17, 2020, about 2.2 million cases and more than 150,000 deaths have been reported worldwide [37, 20]. First cases in Germany were reported at the end of January 2020. As of April 17, 2020, the Robert Koch-Institute (short: RKI) counts over 130,000 confirmed infections and about 4,000 deaths in Germany.

Due to the novelty of the virus, it is reasonable to assume that nobody has prior immunity, that is, the entire human population is potentially susceptible to SARS-CoV-2 infection [11]. Droplet transmission, occurring when a susceptible person comes in close contact with an infective who has respiratory symptoms (coughing and sneezing), seems to be the main infection route of SARS-CoV-2 in the population. Contact transmission, via surfaces in the immediate environment or objects used by an infectious person is also possible [29, 36].

After an incubation period which varies from 2 to 14 days, first unspecific symptoms, such as fever, cough, sore throat or muscular pains, might appear. The illness can then become more acute and lead to difficulties in breathing or progress into severe pneumonia. In critical cases, multi-organ failure can follow and lead to death [36].

Chinese data on the first two months of the outbreak report that about 80% of observed cases are mild to moderate infections [30]. According to the distribution of reported cases in early April in Italy [13], about one third are either completely asymptomatic (7%), pauci-symptomatic (15%) or non-specific infections (13%), about 41% are mild respiratory infections which do not require hospitalization, whereas about 20% showed severe symptoms requiring hospitalisation (20%), and about 3% of the reported cases required intensive care. Severe illness and death are more common among the elderly or patients with other chronic underlying conditions [14].

Whether or not asymptomatic infections do contribute to the spread of the virus is yet to be clarified [36]. One study on the first COVID-19 cases near Munich reports that the infection appeared to be transmitted during the incubation period of the patients, and suggests to reconsider the role of asymptomatic persons in the transmission dynamics of the current outbreak [28], as does another study of patients in Guangzhou Eighth People’s Hospital [17].

There is to date no specific treatment nor vaccine against SARS-CoV-2 [11]. Nevertheless, first studies showed that people infected with the virus develop specific antibodies [39], and based on previous knowledge of other Coronaviruses, like SARS or MERS, it is possible that also SARS-CoV-2-specific immunity will last for about three years [30].

Since January 2020, several mathematical models for understanding and controlling the spread of SARS-CoV-2 have been proposed, in particular concerning the Chinese outbreak [24, 27, 33, 34, 25] and the risk for this to spread to other countries [5, 21]. In all affected regions, non-pharmaceutical interventions to reduce virus transmission were introduced. Mathematical methods were developed to study the impact of such interventions [19] and simulate the effects of relaxing control measures [26]. Possible scenarios for the spread of SARS-CoV-2 in Germany were considered in preliminary studies by an der Heiden und Buchholz [2] who projected the effect of various model parameters (both related to unknown properties of the pathogen and the disease, and to the effect of control measures) on the number of cases, in particular of those requiring hospitalization and intensive care, as well as deaths. Among the key parameters to be approximated by a mathematical approach is the so-called basic reproduction number (ℛ_0_), a metric which indicates the average number of secondary infections generated in a fully susceptible population by one infectious individual over the course of the infection. The basic reproduction number is a reference parameter in mathematical epidemiology used to understand if, and in which proportion, a disease will spread among the population. In simple disease transmission models, ℛ_0_ *>* 1 implies that the virus will spread in the population, and the larger the value of ℛ_0_, the harder it is to control the epidemic. First studies on the initial outbreak in China estimated that the basic reproduction number of COVID-19 could range between 2.2 [24, 27] and 4.71 [33], or could even be larger than 6 [34]. The *(effective) reproduction number* ℛ for a given population is the corresponding time-dependent quantity which reflects the number of secondary infections generated by one infectious individual in the current population, and is affected by intervention measures aimed at controlling the spread of the disease. Among the first studies to asses the practical implications of public health interventions, Tang et al. [34] identified contact tracing followed by quarantine and isolation, as well as travel restrictions as the most effective measures to contain the epidemic. Meyer-Hermann and coauthors [22] recently predicted the evolution of the reproduction number for the spread in Germany, with detailed analysis for all federal states and observed that as of April 3, 2020 the reproduction number ℛ was lowered to values near 1 in all federal states.

In this study, we predict the spread of COVID-19 among the German population by means of mathematical modeling, simulating the implementation or withdrawal of non-pharmaceutical interventions. First model results were presented in our preliminary work [4]. Here we present a follow-up of our study, including most recent data and information about testing activity made available by the RKI [32]. The proposed setting allows to investigate how a specific intervention scenario affects the dynamics of the epidemic, with particular attention to interactions between individuals of the same or different age groups (children, adults, and people older than 60 years). We consider the following scenarios:

- **Minimal intervention**: The main factor slowing down disease transmission is people’s increased awareness in response to initial recommendations coming from health institutions and local governments, and to media coverage (e.g. washing hands, proper coughing and sneezing, keeping distance from obviously sick persons, limited (self-)quarantine of known or suspected cases);
- **Baseline scenario:** Hitherto adopted main control measures (closure of schools and universities, remote working policy, isolation of identified cases, contact restrictions, a partial economic shutdown, and levels of testing activity as of March 15, 2020) are assumed to be maintained throughout 2020. Model parameters which include such control measures were calibrated on reported cases in Germany as of April 5, 2020 and were used to project data until April 14, 2020;
- **High vigilance**: This scenario is obtained by enriching the *baseline* measures with significantly increased testing activity (not only suspected SARS-CoV-2 infectives but also persons without symptoms or known close contacts to identified cases) and a strict isolation protocol of detected SARS-CoV-2 cases for about two weeks;
- **Educational/economic reopening**: Partial lifting of the restrictions imposed thus far, gradually reopening schools, universities, and resuming economic activities, though largely maintaining remote working policy and limiting use of public transport service and organized club activities. Significantly increased testing activity and strict isolation protocol of detected SARS-CoV-2 cases are maintained;
- **Phase-out**: Gradual lifting of most control measures applied so far (reopening schools, resuming work, regular public transport service, resuming most social and economic activities). Increased testing activity and strict isolation protocol of detected SARS-CoV-2 cases are maintained, and the population is assumed to uphold minimal awareness measures;
- **Cautious phase-out**: As the *phase-out* scenario, but with slower rollback to educational, social and economical activities, and with improved measures to protect elderly and people at risk.

Our model predicts that the current control measures are necessary to slow down or even suppress the spread of the epidemic, and that the removal of restrictions in favour of social and economic activities will accelerate the growth of case numbers unless it is accompanied by a significantly strengthened testing and case isolation policy. However, under such increased vigilance, combined with particular care regarding patients at high risk, we project that a gradual phase out of the most severe measures starting around May 5 will lead to a progression of the epidemic that is sufficiently slow to be handled by the health care system.

## 2 Methods

### 2.1 Mathematical Models

The mathematical models adopted for this study are based on systems of differential equations that describe interactions between different groups of individuals in the population. The proposed approach extends the known S-E-I-R (susceptibles-exposed-infected-recovered) model for disease dynamics [6]. Individuals are classified according to their status with respect to the virus spread in the community. In particular we distinguish between individuals who have been exposed to the virus but are not yet infectious, asymptomatic infectives, infectives with mild or influenza-like symptoms (not reported as SARS-CoV-2 infections), and reported SARS-CoV-2 infectives. We assume that infected patients without SARS-CoV-2 diagnosis are unlikely to die of the virus-induced disease. Undetected infections lead to undetected recoveries in the population, which cannot be reported unless testing for ongoing (virus detection) or previous (antibody detection) SARS-CoV-2 infections is performed.

#### 2.1.1 The core model - homogeneous population

The simplest approach that we adopted to understand the evolution of the epidemic in time is based on the assumption that the population is homogeneous (in particular with respect to space, as well as to age and social habits of individuals). Of course, this simplistic assumption does not reflect the multidimensional complexity of the ongoing situation, but it can help in determining major factors affecting disease spread.

Individuals are classified as follows. Susceptible individuals (*S*) are those who can be infected with SARS-CoV-2 virus. Exposed individuals (*E*) have been infected with SARS-CoV-2 virus, though they are not yet infectious, nor symptomatic. After a latent phase (on average about 5.5 days from infection [30]) the first COVID-19 symptoms appear (with probability *ρ*_0_) and the exposed individual becomes infectious. We distinguish between asymptomatic undetected (*U*) COVID-19 patients, symptomatic but not yet detected infectives (*I*) and reported COVID-19 cases (*H*). For individuals developing symptoms, the probability of the disease being detected immediately is *η*_0_, the probability of detection at a later stage is denoted by *η*_1_. Individuals who recovered from a detected (*R*) or an undetected (*R*_*u*_) infection, as well as patients who died from the infection (*D*), are removed from the chain of transmission. The transmission diagram of the model is depicted in Fig. 1. Susceptible individuals can be infected via contacts with asymptomatic (transmission rate *β*_*U*_), symptomatic undetected (*β*_*I*_) and reported cases (*β*_*H*_). We assume that asymptomatic infectives do not restrict their contacts to others, and therefore have higher transmission rates than symptomatic infected individuals. Detected cases are supposed to reduce their contacts even further. Due to limitations in the identifiability of the parameters with the available data, we fix the ratio between *β*_*I*_, respectively *β*_*H*_ and *β*_*U*_ and estimate the latter. Duration of latency (1*/γ*_*E*_) and infectious periods (1*/γ*_*I*_, 1*/γ*_*H*_ and 1*/γ*_*U*_, respectively) are assumed in accordance with available literature. Further details on parameter assumptions are given in Table 1 and the model equations are given in the appendix.

**Table 1:** Main model parameters in the core model. Parameter descriptions, values and references.

|  | Description | Value [unit] (Refs.) |
| --- | --- | --- |
| $\beta_U$ | Transmission rate of asymptomatic undetected infectives | fitted [1/(d·individual)] |
| $\beta_I$ | Transmission rate of symptomatic undetected infectives | $0.8\beta_U$ (assumed) |
| $\beta_H$ | Transmission rate of detected infectives | $0.1\beta_U$ (assumed) |
| $1/\gamma_E$ | Mean incubation period | 5.5 [d] [3, 31, 2] |
| $1/\gamma_I, 1/\gamma_H$ | Mean duration of symptomatic infection | 7 [d] [22, 23, 30] |
| $1/\gamma_U$ | Mean duration of asymptomatic infection | 6 [d] [23, 30] |
| $\delta_H$ | Case mortality of undetected infections | 0.057 [14, 22] |
| $\rho_0$ | Probability of developing symptoms | 67% [13] |
| $\eta_0$ | Probability of detection during latency | fitted |
| $\hat{\eta}_1$ | Probability of detection while asymptomatic | 7% (assumed) |
| $\eta_1$ | Probability of detection while symptomatic | fitted |

**Figure 1:**
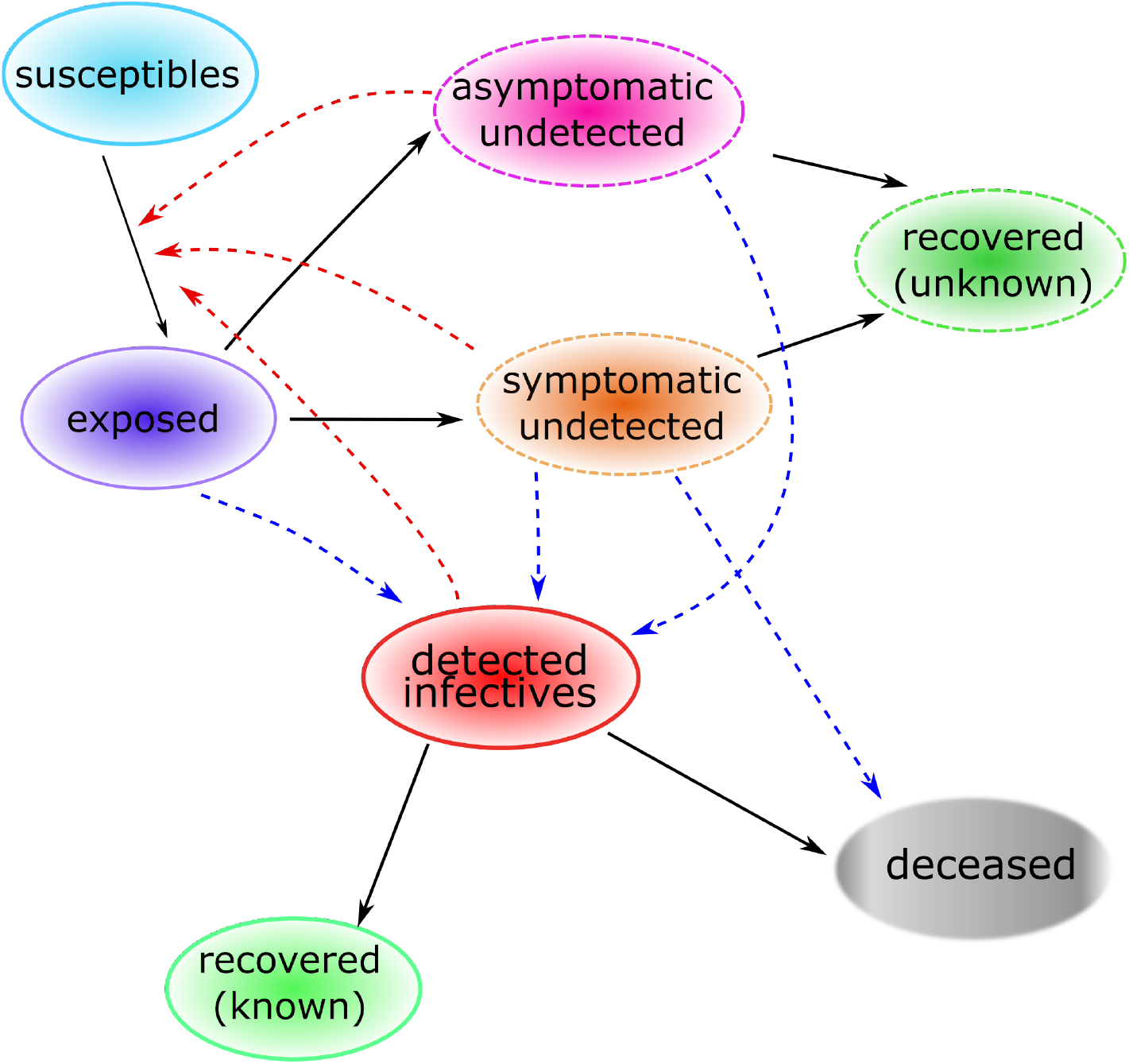
Core model structure for COVID-19 outbreak in Germany. Solid arrows indicate transition from one compartment to another, red dashed arrows indicate virus transmission due to contact with infectives, blue dashed arrows indicate detection of infections due to testing activities. Upon infection with SARS-CoV-2, susceptible individuals enter a latent phase, in which they are not yet infectious nor symptomatic. After the latent phase, individuals become infectious, may develop symptoms and may be detected as COVID-19 cases. Infected individuals who recovered from a detected or an undetected infection, as well as patients who died upon infections, are removed from the chain of transmission.

To determine model parameters which could not be inferred from the literature, we use the sum of reported cases from three different time periods (February 28 to March 11, March 11 to March 22, and March 22 to April 11) and fit the data to the sum of H, R, and D using the trust region reflective method as implemented in SciPy [35]. The first period does not include any global contact reducing measures except for school closings in the county of Heinsberg. The second period includes nationwide school closings and an increased use of working from home where possible. The third period includes the closing of most stores and general constraints on contacts in the public sphere. For each periods, we fit *η*_1_, 1 *− η*_0_, and *β*_*U*_, while keeping *β*_*I*_ and *β*_*H*_ proportional to *β*_*U*_ with proportionality factors given in Table 1. To test the sensitivity of the fits to variations in the fit parameters, we add random sampling of the parameters and weigh each sample by the Akaike weight of the generated fit. The Akaike weight is determined based on the Akaike information criterion [1]. For few data points the corrected Akaike information criterion (AICc) [18] takes the form

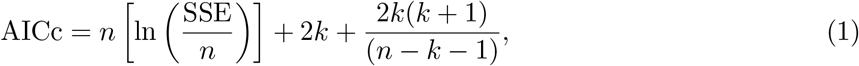

where SSE is the sum of squared errors, *n* is the number of data points, and *k* is the number of degrees of freedom. The weight of each fit *i* in a set of *J* fits is then given as

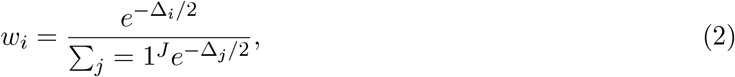

where Δ_*i*_ = AICc_*i*_ *−* AICc_min_. Based on the sample values and their weights, we can construct a histogram for each parameter and derived properties (c.f. Fig. 2). A narrow peaked histogram indicates a well determined parameter with a small standard deviation and a corresponding narrow confidence interval, whereas a flat histogram shows that the parameter may be indeterminate and that the fit is not sensitive to its variations. For a sufficiently large number of points, we can determine, for example, a 95% confidence interval directly from the histogram by excluding the left- and rightmost 2.5% (marked in light blue in Fig. 2).

**Figure 2:**
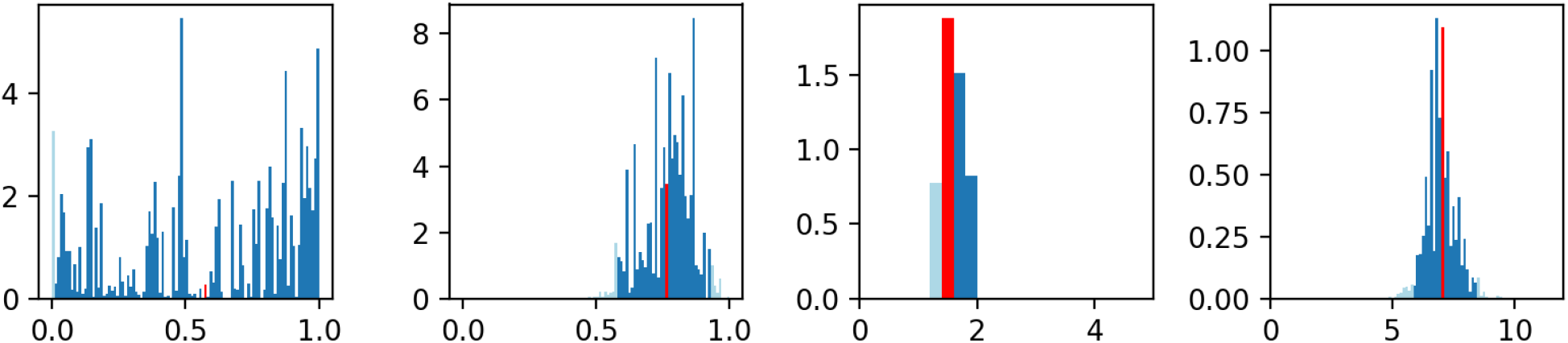
Weighted histograms of *η*_1_, 1*−η*_0_, *β*_*U*_, and ℛ (from left to right) based on stochastic variation of *η*_1_, 1*− η*_0_, and *β*_*U*_. Each sample contributes with its Akaike weight, which is based on a fit to the data from February 28 to March 11, 2020, to the histogram (see Eq. (2)). Dark blue bins lie within the 95% confidence interval, light blue bins lie outside. Apparently, *η*_1_ cannot be determined using these data. The red bin indicates the mean value (of 1 *− η*_0_ = 0.77 (0.58–0.92), of *β*_*U*_ = 1.59 (1.36–1.89), and of ℛ = 6.99 (5.84–8.35)).

#### 2.1.2 Sensitivity analysis of ℛ and peak cases in the core model

Sensitivity analysis of the reproduction number and of the number of cases at the outbreak peak was further investigated by means of scatter plots (results not shown here) and Sobol analysis (SALib library [16] in Python). The Sobol method is a variance-based method for sensitivity analysis that decomposes the output variance into the contributions associated with each input factor. We computed the sensitivity indices of the parameters that could not be determined from literature or are expected to vary between populations and time, in particular the transmission parameters, the probability of developing symptoms and the probability of being detected. Of these parameters *ρ*_0_, *η*_0_, and *η*_1_ are varied between 0 and 1, *β*_*U*_ is varied between 0 and 3, and *β*_*I*_ and *β*_*H*_ are assumed to be fixed multiples of *β*_*U*_ as detailed in table 1. All the parameters are varied simultaneously to also observe their interaction effects. The first-order Sobol sensitivity indices (S1), which describe the contribution to the total variance due to each parameter alone [38], indicate that *β*_*U*_ has the expected very strong effect on the calculated (cf. the appendix) reproduction number (*S*1(*β*_*U*_) = 0.868542*±*0.069355). Also the probability of developing symptoms in early stage has an effect on ℛ, but this is much smaller than the effect of *β*_*U*_ (*S*1(*ρ*_0_) = 0.041441 ± 0.023002). Early-stage detection also has a similar small impact (*S*1(*η*_0_) = 0.051242 ± 0.026127), while the probability of detection while symptomatic has an even lower impact (*S*1(*η*_1_) = 0.001928 ± 0.004873). Both the transmission rate of asymptomatic individuals and the probability of developing symptoms in early stage seem to have strong effects on the maximum number of active reported infections (*S*1(*β*_*U*_) = 0.319293 ± 0.048579, *S*1(*ρ*_0_) = 0.422847 ± 0.058169). Detection probabilities also have a small impact on the maximum number of active infections reported (*S*1(*η*_0_) = 0.047637 ± 0.030934, *S*1(*η*_1_) = 0.050986 ± 0.029052).

#### 2.1.3 Population structured by age and stages of infection

Refining the basic model described in Sec. 2.1.1, we include age groups and stages within infective compartments. This allows to consider features like the immune response of individuals during infection or social behavior, in particular interactions among individuals of the same or different age groups. Based on the statistical analysis of the RKI data in section 4, we distinguish three groups: children (0-14y), adults (15-59y) and people 60y or older. For each age group the model tracks susceptibles (*S*), exposed (*E*), asymptomatic infectives (*U*), (undetected) symptomatic infectives (*I*), diagnosed infectives (*H*), recovered (*R*), undetected recovered (*R*_*u*_), and deceased (*D*) individuals. To obtain more realistic distributions for the duration of the exposed and infective compartments, we split each of these in three stages (*E*_*j*_, *U*_*j*_, *I*_*j*_ etc. *j* = 1, 2, 3). This is a classical extension of the standard disease transmission model to account for non-exponential distributions of incubation and infectious periods (cf., e.g., [5] for another example of application in modeling COVID-19). This results in a total of one plus nine infective compartments per age class (stage *E*_3_ is assumed to be infective as well, as individuals have been reported to be infectious before symptoms onset [17, 30]). The age classes evolve in parallel (maturation during the course of the outbreak is neglected), but are coupled to one another by contact rates and disease transmission among individuals of different age groups.

After virus transmission, an exposed individual is assumed to travel through the stages *E*_1_, *E*_2_, and *E*_3_ before entering either of the infection stages *I*_1_, *U*_1_, or *H*_1_. In the course of the disease the individual will then pass through the infection stages of the respective compartment. Both symptomatic and asymptomatic infectives in stages *U*_*i*_ or *I*_*i*_ can enter stages *H*_*i*+1_ or *H*_*i*_, respectively, by being tested. The probability of being tested and positively diagnosed is assumed to be larger for symptomatic than for asymptomatic individuals. Assuming the chance of late symptom onset to be covered by the nonzero chance of a longer incubation period, we have neglected any conceivable transitions from *U* to *I*. The individual leaves the last stage of infection (*U*_3_, *I*_3_) by either recovering to *R* or *R*_*u*_, depending on whether the infection has been diagnosed. Given the current efforts to detect and control the spread of the disease, it seems a justified assumption that the vast majority of deaths caused by SARS-CoV2 are investigated, hence we assume only diagnosed individuals (*H*) to be fatally affected by the disease.

In this structured model we can include control measures which explicitly affect different age groups. In particular we consider (i) general increased awareness in the population due to the effect of media (M), as well as (ii) active control due to main intervention measures adopted throughout Germany since March 2020. These control measures include: (CS) Closure of all schools, universities, sports clubs and canceling public events; (HO) reduced contacts in workplaces and outside the household (restaurants, bars, public transport); (T0) initial efforts to improve detection by more testing; (IC0) isolation of infected cases; and lock-down measures (LD) closing most activities and prohibiting more than two people gatherings in effect by the end of March. Details are summarized in Table 2.

**Table 2:** Summary of control measures for mitigation of COVID-19 infections adopted in Germany as of April 4, 2020. The indicated percentages are meant to be compared to the state without the respective measure (i.e., the whole has a multiplicative effect).

| Label | Policy | Description/Working assumptions |
| --- | --- | --- |
| CS | Closure of all schools, universities, sport clubs. Public events are canceled. Stricter social distancing encouraged | Overall reduction of activities, decreasing all entries by 10%. Moreover, a strong effect on the shape matrix reducing child-child contacts by 80%, adult-adult contacts by 20%, and senior-senior by 10%. Adopted nationally from March 14 (day 28), fully in force March 16 (day 30). |
| HO | Remote working policy (home office), closure of all restaurants and bars, reduced (use of) public transport service. | Reducing contacts: child-child (- 20%), child-adult (- 20%), adult-adult (- 50%), child-senior (- 30%), adult-senior (-30%), senior-senior (- 30%). Assumed to be applied at national scale from March 13 (day 27), contact reductions fully achieved after 13 days. |
| M | Awareness raising due to media, canceling big events | Information and media activities increase social distancing and personal hygiene (e.g., washing hands), starting on February 25 (day 10). Contacts are on average reduced by 20%, achieved by activity reduction (higher relative decrease for seniors than for juniors and adults). |
| T0 | Testing | Increased testing activity starting February 28 (day 13), most significant increase between March 9 and March 15 (days 23 through 29) Approx. 10-fold increase in detection rates compared to previous levels. |
| IC0 | Isolation of infected cases | Identified and suspected infected cases (self) quarantined for 2 weeks. Reduces *effective* contact rates (for individuals in specific groups by up to 40%); implemented starting on February 28 (day 13), in effect by March 7 (day 21). |
| LD | Partial economic and social activities | Closing non-critical businesses, gatherings of more than two or three persons sanctioned, decreases contact rates by another average 30%, starting March 22 (day 36), in full effect by March 27 (day 41). |

Reported cases for the three age groups were used to fit the model including control measures as indicated in Table 2, until April 5 (Fig. 3). The obtained setting was used to predict data from April 6 to April 14, as well as long term predictions of the *baseline* (BSL) scenario, in which the currently applied control measures (CS, HS, TO, ICO and LD) and the awareness due to the effect of media are maintained until the end of the year (Fig. 7b). The *baseline* scenario has been then used to simulate different possible future scenarios (cf. Table 3). On the one hand, we have enriched the current control measures by further increased testing activity (T1+), stricter isolation of known cases (IC) and increased social distancing from individuals at risk (IO). On the other hand, we have considered possible rollback scenarios (cf. Sec. 3).

**Table 3:** Possible measures included in simulated scenarios, starting from April 7. Some of them are by now in effect.

| Label | Policy | Description/Working assumptions |
| --- | --- | --- |
| T1+ | Significant further increase of testing | Further increase in testing by at least four-fold, not only of individuals with relevant symptoms and/or contact to identified cases but also of asymptomatic persons with any suspicion of being infected; starting on day 60 (April 15). |
| RB | (partial) Rollback of measures<br>HO, CS, and/or LD<br>CS:<br>HO:<br>LD: | Gradually taking back the effects of contact rate reductions according to a possible schedule<br>(partially) reopening schools and child care facilities from May 4 (day 79), reduces effect, particularly on children, by approx 70%, reopening universities etc. on May 18 (day 93), reopening clubs and social activities on June 1 (day 107)<br>restricting HO policy to cases where easily applicable and cautiously reopening restaurants decreases the effect by approx. 60% by May 18 (day 93), final return to full activity by June 1 (day 107)<br>Lifting most severe curfew-like restrictions on May 4 (day 79) reduces effect by 50%, gradual further reductions on May 18 and June 1 to pre-measure level |
| IC | Stricter isolation of known infectives compared to IC0 | Reduces activities rates of identified SARS-CoV-2 infectives by another -50% (compared to IC0), starting on April 10 (day 55), in full effect by April 20 (day 65). |
| IO | General social distancing (incl. self-separation) of individuals at risk, in particular elderly | Includes particular hygienic care when dealing with risk group individuals, reduction of senior susceptibility by another 40%, starting on April 7 (day 52), in full effect on April 10 (day 55) |

**Figure 3:**
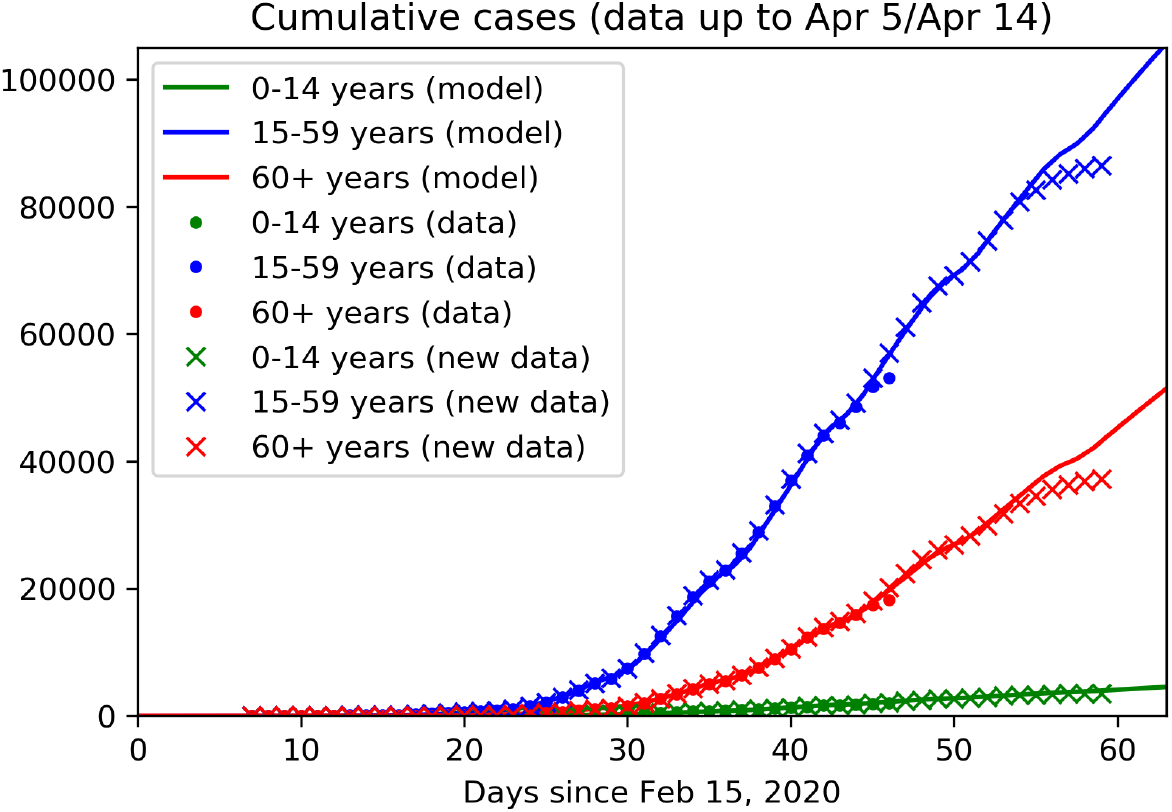
Age and stage structured model: Data fit and predictions. Continuous curves show model solutions, dots are reported data up to March 25 (as used in our previous work [4]), cross denotes reported data as of April 15. The model is calibrated on collected data up to April 5 (day 50), data from April 6 to 14 are used for validation. Colors denote the three different age groups: juveniles (0-14 y, green), adults (15-59, blue) and seniors (60y and older, blue). It should be noted that the most recent data tend to be lower than expected since not all cases detected on these days have been reported to the RKI, yet.

Control measures have the fundamental effect to reduce contacts between individuals, henceforth transmission of the virus from person to person. In our modeling assumptions, we consider contacts in three different realms: at home, at work and school, and in other locations, most notably during leisure activities. For each activity, we assume specific contact distributions depending on the ages of the individuals. For example, we assume high contact rates among adults and between adults and children at home. The separation of the realms allows for each intervention to produce a specific effect. For example, school closures predominantly act on the child-child contacts in the school realm. An example of how control measures reduce the transmission rates in the *baseline* scenario is illustrated in Fig. 4a. In contrast, in rollback scenarios a partial lift of control measures leads to increased transmission rates (Fig. 4b).

**Figure 4:**
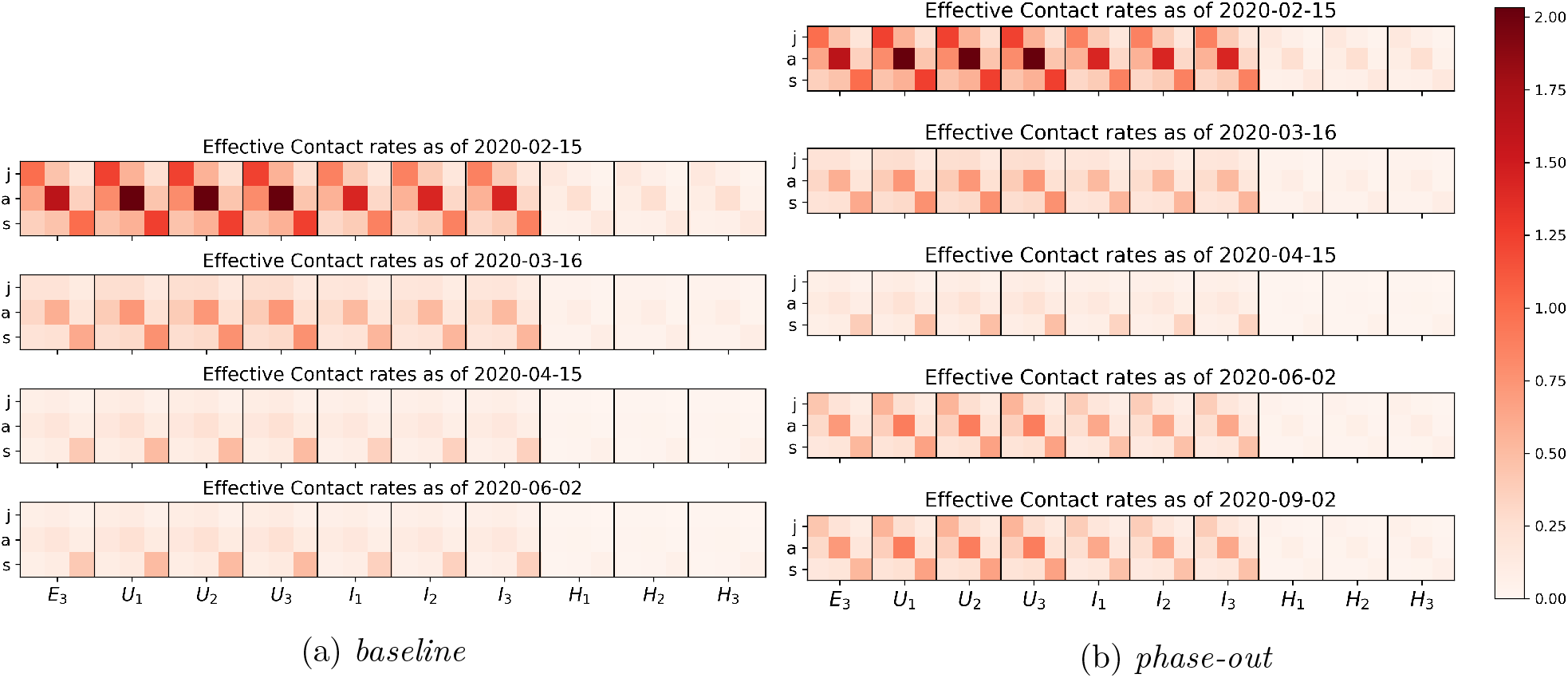
The evolution in time of contacts between juvenile (j), adult (a) and senior (s) susceptible individuals (columns) and infectious individuals in the late latent phase (*E*_3_) or one of the infected phases (*I*_1_, … *H*_3_). (a) In *baseline* scenario control measures will be maintained over time hence contacts and transmission of the virus will remain low, whereas (b) a lift of all control measures (*phase-out* scenario) will slowly lead to a new increase in contacts and virus transmission.

## 3 Results

In the core (non-structured) model we estimated how contacts, hence the reproduction number, decreased in the three considered time slots since the beginning of the epidemic using fits to the data. The left panel of Fig. 5 shows the fits on top of the data. Each period is described by a different value for the reproduction number: ℛ = 6.99 for the first period, ℛ = 3.79 for the second period, and ℛ = 1.04 for the third period. This clearly shows the success of the contact reducing measures. The inset in Fig. 5 shows the predictions based on the three fits until mid May showing the catastrophic increase of infections that would have resulted if no or only insufficient measures had been taken. The right panel shows a range for cases requiring hospitalization and intensive care using the latest prediction of the reproduction number. Under the current prediction these numbers could be handled by German hospitals. Though this is an indicative estimate only, it clearly shows that the reproduction number consequently decreased from ℛ = 7 at the end of February to almost ℛ = 1 at the beginning of April.

**Figure 5:**
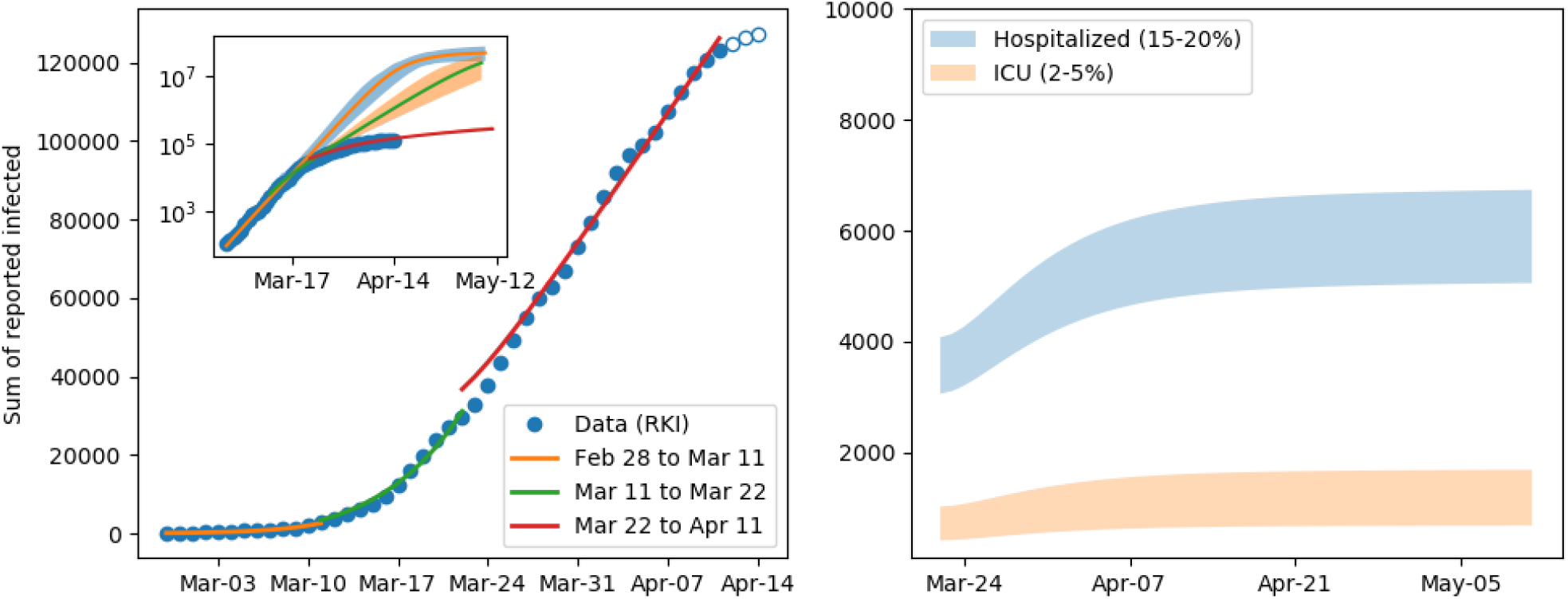
Model fit results and extrapolation. The left panel shows the fits in the 3 periods, clearly indicating the ability of the model to capture the dynamics. The open circles are data points not used for the fit. Due to the inherent latency of the reporting in Germany, these points are not reliable, yet. The inset shows in addition the prediction for the sum of reported cases. Without intervention the number of infections could have surpassed ten million cases by mid April. The bands around the lines indicate the 95% confidence level of the fit based on stochastic variation of the parameters described in 2.1.1. The right plot shows the number of individuals that required hospitalization either in the low care ward or ICU.

In the *baseline* scenario for the refined age and stag e structured model, where control measures apply continuously over time, we observe that the dominant eigenvalue of the linearization of the system about the disease-free equilibrium (DFE, i.e., a completely susceptible population) decreases in time and eventually crosses zero (Fig. 6a). This corresponds to the reproduction number dropping below ℛ = 1 (cf. [10]). The fluctuations which we observe in Fig. 6a are due to what we call the *weekend-effect*, namely the weekly fluctuation observed in reported cases (cf. Fig. 6b).

**Figure 6:**
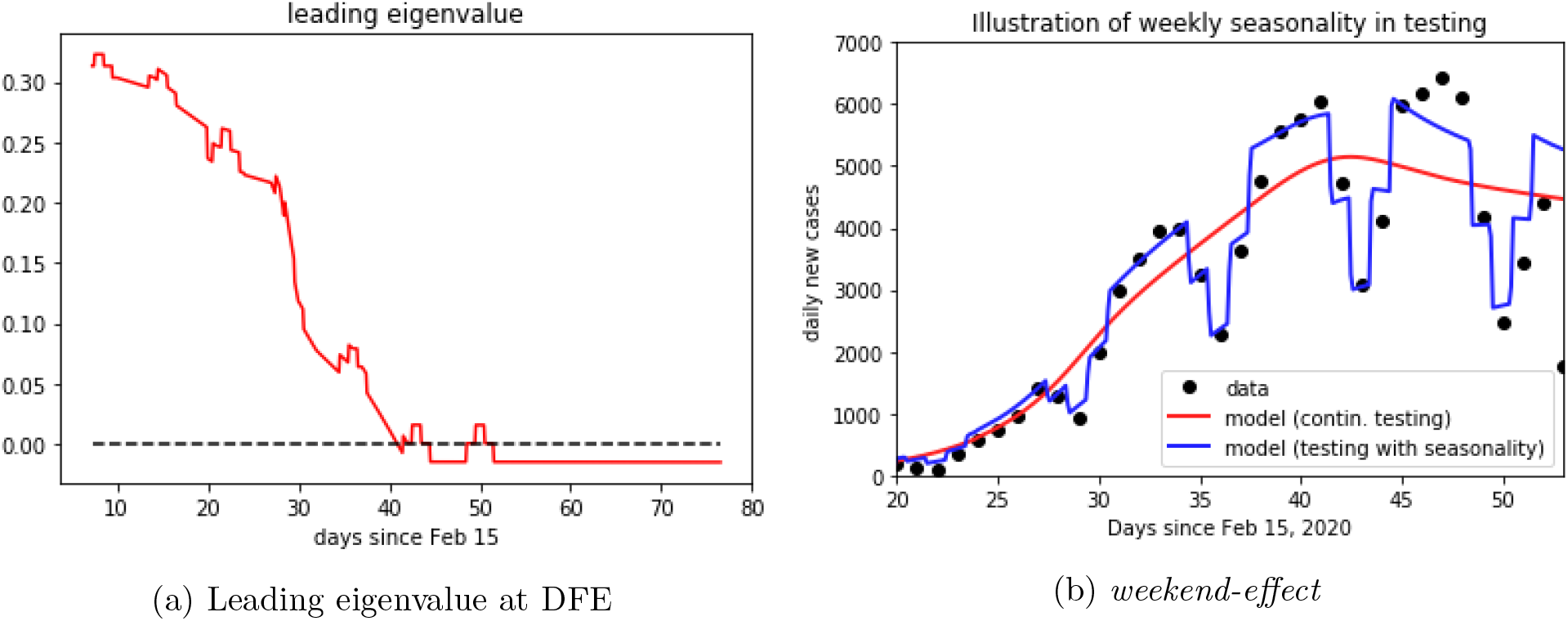
(a) Evolution in time of the dominant eigenvalue of linearization of the age and stage structured model at the disease free equilibrium (DFE). The dominant eigenvalue crossing the zero axis corresponds to the reproduction number crossing the threshold value 1. (b) The *weekend effect* accounting for fluctuations in case detection and reporting. Black dots denote daily reported new cases, continuous curves show the model solutions with (blue) and without (red) time-depending testing rates.

Let us now discuss results on the simulated scenarios in detail. In the *minimal intervention* scenario, we assume that no specific measures to reduce contacts between individuals (school closures, interruption of most social and economical activities) nor increased testing activity were undertaken. Under these assumptions, the initial rapid increase of cases would have gone on unabatedly, and the number of infected individuals requiring hospitalization or even intensive care would have reached unmanageable levels within weeks. This scenario is purely counterfactual and is only detailed here to evaluate the effects of the measures adopted so far. Model simulations (Fig. 7a) show that in this scenario a peak in infections would have been reached at the beginning of May 2020 (day 79 since February 15), with about 12 million active infections on the peak day. Over the course of the infection about 75 million people would have been infected and 1.6 million would have died. Fortunately, taking into account interventions adopted so far, the actual course of the epidemic is less dramatic, and the model simulations predict a significantly better outcome.

**Figure 7:**
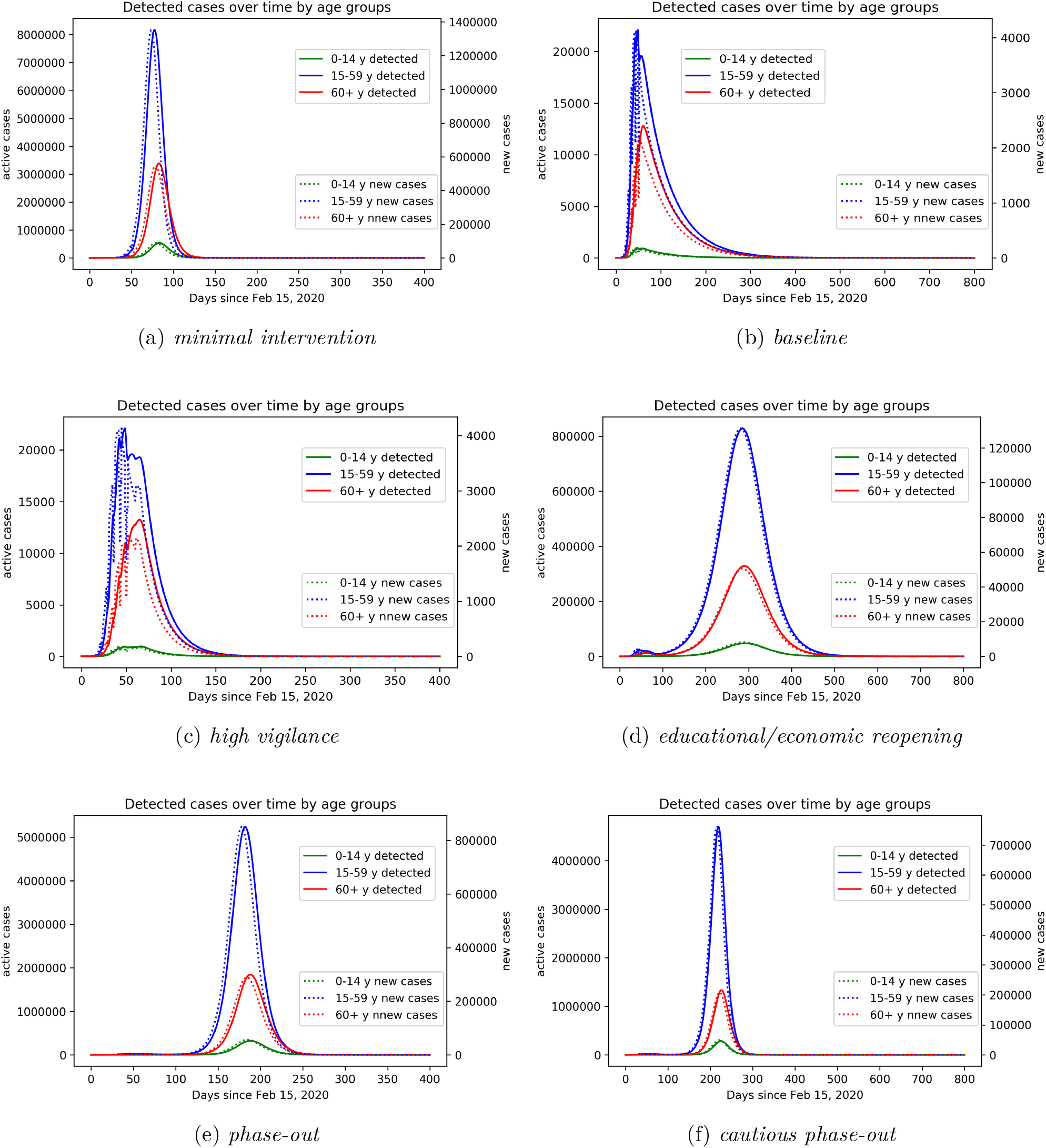
Model prediction for simulated scenarios. (a) *minimal intervention*: increased awareness, quarantine of known or suspected cases, testing of patients with symptoms and contact history; (b) *baseline* scenario: *minimal intervention* scenario increased with school closure, high reduction in economical activities, contact limitation, high testing activity; (c) *high vigilance*: *baseline* scenario enriched by isolation of detected cases, combined with increased testing activity; (d) *educational/economic reopening*: reintroducing in three phases contacts at schools, workplaces, public transportation service; (e) *phase-out*: rollback of all introduced control measures, up to minimal interventions, accompanied by increased testing also of asymptomatic individuals and strict isolation of identified cases; (f) *cautious phase-out*: similar to (e), but with slower rollback to regular activities, accompanied by strongly increased testing also of asymptomatic individuals, strict isolation of identified cases, and reduced contacts with elderly and risk groups.

Compared to our initial investigations [4], here we have adjusted the *baseline scenario* by taking into account the further restrictive measures adopted in the last week of March 2020 (reduction of economic activity, restrictions on meetings in public space, and further increased remote working activity). Said enhanced intervention scenario suggests that the number of active cases peaked with about 33,000 infected individuals at the beginning of April 2020 (Fig. 7b). This is in accordance with recent modeling studies [22], which suggest that the effective reproduction number ℛ *≈* 1 as of April 2, meaning that the disease free equilibrium is on the verge of instability (cf. Fig. 6a)^1^. This makes the predictions for this scenario particularly sensitive to assumptions about the model parameters as small variations of ℛ make a striking qualitative difference between further exponentially rising case numbers or a slowdown of the epidemic. Taking the latest case numbers (up to April 8) into consideration and assuming that in the weeks to come the testing activity does not take significant slumps on weekends anymore (cf. Fig. 6b), we project the number of infected individuals, both detected and undetected, to decrease over the coming months. The total number of fatalities would be reduced by more than 90% as compared to the *minimal intervention* scenario (Fig. 8d), and the capacity of the health care system would not be severely challenged (see also Fig. 8b).

**Figure 8:**
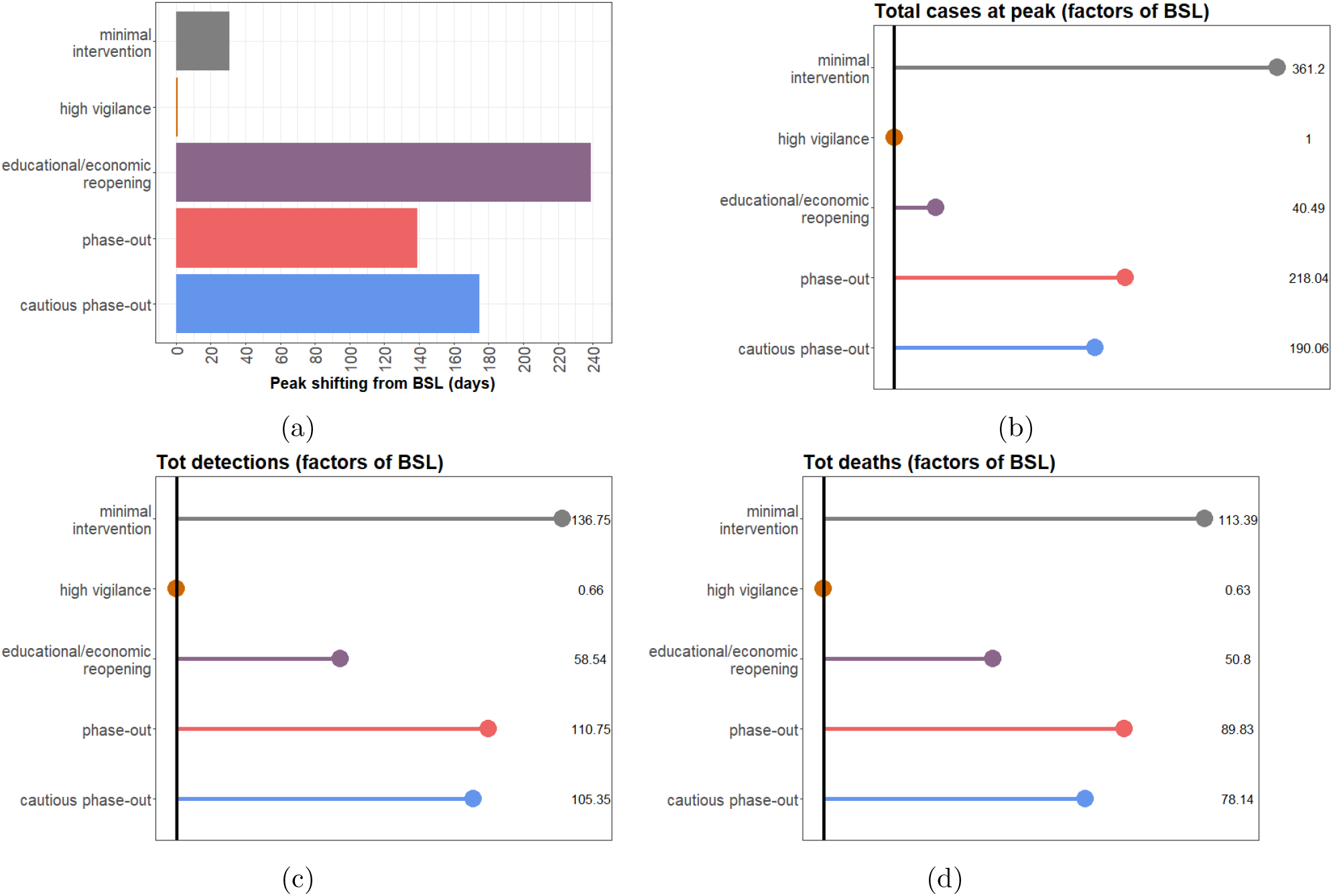
Statistical comparison of model output for the *baseline* (BSL) scenario and the considered possible alternatives. (a) Peak shifting (in days) compared to BSL; (b) Differences in reported cases (factor) at the day of the peak; (c) Differences in total detected cases (factor); (d) Differences in total deaths (factor). For all rollback scenarios, results refer to the second peak of the outbreak.

In the reported data, individuals are counted as “recovered” if they are no longer symptomatic and 14 days (supposedly the longest infectious period under normal circumstances) have passed since the positive test. In contrast, the *average* infectious period, as chosen for the simulation, is about 1*/γ*_*I*_ = 7 *d*, meaning that individuals on average recover seven days after becoming infectious. Therefore, the number of recovered individuals according to the model is higher than the officially recorded figure, and that in turn leads to lower numbers of active reported cases in the model as compared to the official data. Using data up to April 5 to estimate parameters, the model predicts 116,000 cumulative reported cases for April 9, and about 149,000 cases for April 16. Maintaining the *baseline* measures throughout the year (we are assuming no seasonality of the disease) would lead to the eradication of the epidemic. Simulations of this scenario (Fig. 7b) suggest about 550,000 infections (about 185,000 thereof asymptomatic), and 14,000 deaths over the course of the epidemic. Enriching the *baseline* scenario with further increased testing activities and even stricter isolation of detected and suspected cases (*high vigilance* scenario, Fig. 7c) would shorten the time necessary to call the disease eradicated (beginning of September 2020), but widen the peak of active cases. Stricter isolation of confirmed and suspected cases and improved testing activity would further reduce the spread of the virus, forecasting about 361,000 infections (out of which about 108,000 asymptomatic), and 9,000 deaths over the course of the epidemic.

The above scenarios assume that the current restrictions on public life remain in effect over a long period. As this does not seem to be feasible in practice, we consider further scenarios which include an at least partial lifting of the restrictions imposed thus far. In contrast to our previously simulated scenarios [4], here we assume a gradual reopening of economical and educational activity (Fig. 7d). We assume this to start in about three weeks from now (May 4) with reopening schools and childcare facilities as well as many shops, gradually proceeding to reopening universities, restaurants and other economic activities, and finally resuming on-site work and most club activities from June 1 on. Combining this partial rollback with further increased testing activity and isolation of identified cases (*educational/economic reopening* scenario) would lead to a (second) peak in active cases (1.3 million) towards the end of November 2020. If no restrictive measures and interventions were to be (re)introduced, the simulation of the model results in about 32 million total infections and 730,000 deaths over the course of the epidemic, which seems to occur only by the end of the summer 2021 and under the assumption that no reliable treatment becomes available by then.

The last two scenarios that we present here suggest that a complete, though gradual, rollback of all introduced control measures would lead to a second peak towards the end of August 2020 (Fig. 7e, *phase-out* scenario), or end of September 2020 in case of slower reintroduction of regular activities (Fig. 7f, *cautious phase-out*). In both cases, the second peak would be anticipated (Fig. 8a) and the number of infections at this peak would be way larger than in the *educational/economic reopening* scenario (7.2 million and 6.3 million active cases in the *phase-out* and *cautious phase-out* scenarios, respectively; cf. Fig. 8b).

## 4 Discussion

In this work we proposed a mathematical model for predicting the evolution in time of detected COVID-19 infections in Germany, taking in particular into account the age distribution of cases. Distinguishing between people in different age groups the model allows for one thing to better characterize contacts between individuals (e.g. child-child contacts being typically different than senior-adult contacts, cf. [26]), for another thing to fine-tune the effect of intervention measures on contacts reduction (cf. Fig. 4).

Our results indicate that the current measures lead to a significant reduction in the reproduction number, R, which is approximately one in the second week of April (Fig. 6a). This matches with estimations of the RKI [31] and findings by other groups which are also currently studying the situation in Germany [9, 22]. This naturally results in a significant uncertainty of the projection since small deviations of the parameters can make the difference between further growing active case numbers or slowly declining numbers.

We parametrized the model using, if possible, known or previously estimated parameter values, in particular those concerning the evolution of the disease (latency time, duration of infection, death rates), or plausible assumptions (e.g. for the relation between infectivity of detected and undetected infectives) as explained in Sec. 2 and the appendix. Uncertainties of parameters which could not be inferred from the literature or well identified from the data, were investigated via stochastic sampling and Sobol analysis (cf. Sec. 2). Such uncertainties could be reduced by integrating further data in the model, e.g. estimated contacts or estimates for the number of unreported cases. In our *baseline* scenario we assumed the current reporting ratio in Germany to be rather high (over 50% as of mid April). Assuming a lower reporting ratio yields more undetected cases, and therefore leads to a higher estimate of the reproduction number. Analogously, dropping the weekend effect increases the number of active cases at the peak. On the other hand, lower reporting ratios in early April provide a larger margin for improvement by enhanced testing as in our *high vigilance* scenario. The aggressiveness of the virus and hence the mortality among all affected individuals (whether diagnosed or not) is another unknown, but different assumptions about this parameter can be expected to have similar impacts on all the scenarios discussed here. It may be assumed that earlier detection (as in all our scenarios with enhanced testing) and hence earlier and better care for high risk patients may result in a mortality even below the one observed in Germany so far. The limited capacity of the health care system, in particular of intensive care units, was not yet directly considered as a parameter of our refined model. However, the predicted number of infected individuals at its peak can be used as a proxy for the expected demand for health care resources at the height of the epidemic. Assuming that a fixed proportion of infected individuals will require intensive care, the maximal number of infectives for a given scenario directly indicates the maximal load on the health care system for this scenario.

We have conducted simulations covering one year and more starting from the beginning of the epidemic. Some scenarios predict high peaks in active cases and alarmingly high numbers of deaths far into the future. However, these scenarios should only be understood as predictions for the future if no appropriate measures were taken to contain the epidemic. In particular, postponing the date of the peak sufficiently far into the future would provide time for the development of a vaccine or an effective treatment.

## Data Availability

The publicly available dataset provided by the Robert Koch Institute was used for this study.

## Contributions

Conceptualization: MVB; Modelling: JF, MVB; Data curation and analysis: NC; Literature research: SK, HVV, JM, MVB; Calibration: JF, HVV, JM. Numerical simulations: JF, HVV, JM, SK. Writing: MVB, JF, TL, JM, SK.

## Appendix

### Equations of the core model and computation of ℛ_0_

The dynamics of the core model as in Fig. 1 is given by the system

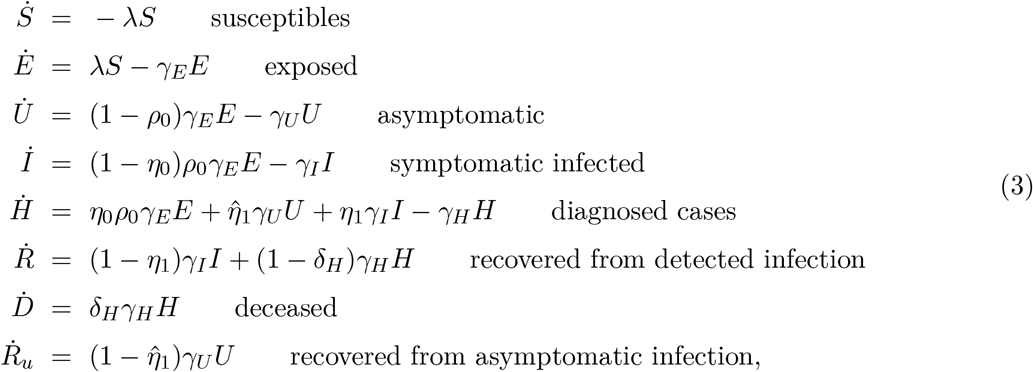

where

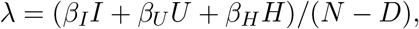

and *N ≈* 83 million is the total population.

As we mentioned in our previous work [4], the basic reproduction number can be calculated analytically, e.g, by means of the next-generation matrix approach [10]. For the above system (3), one obtains

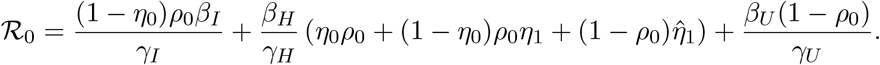

### Equations and parameters of the age structured model

As explained in Sec. 2, we extend the core model (3) by mean of age classes and stages within infective compartments. For each age class *A* we have the following compartments

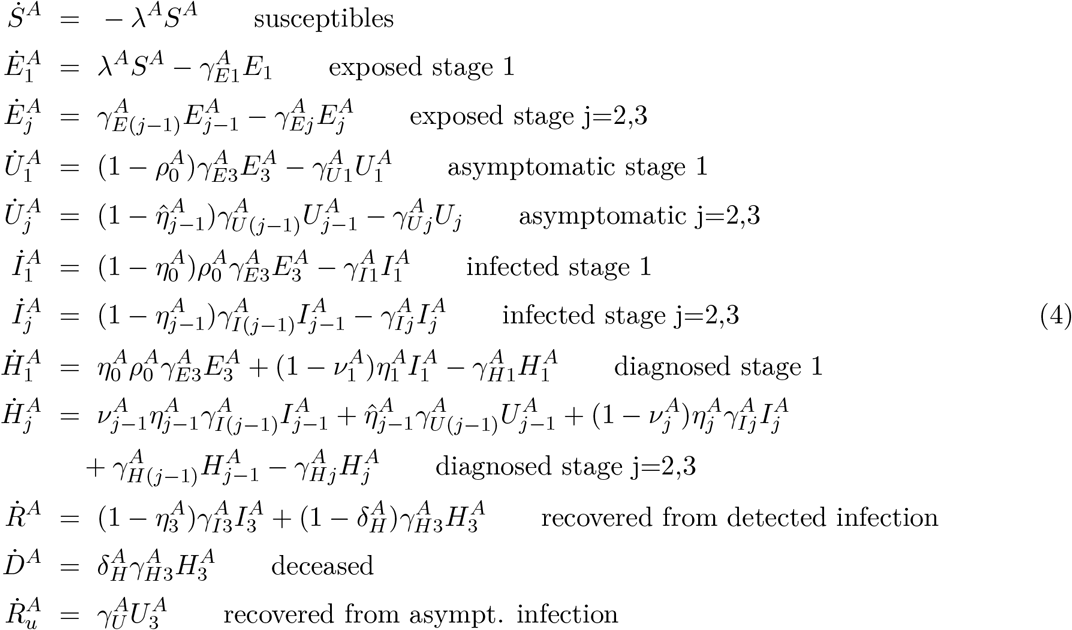

The transmission rate for *S*^*A*^ individuals is given by

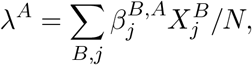

where *N* is the total population (discounted for deaths) and 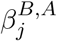 denotes the transmission rate due to contacts of infectious individuals in compartment *X* ∈ *{E, U, I, H}*, age class *B* and infection stage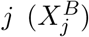 with susceptibles in age class *A*. Contact rates can be summarized in a (3 × 30) matrix 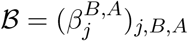.

Entries of the contact matrix *ℬ* are assumed to be influenced by (i) the overall aggressiveness of the virus, *β*_0_, that describes how easily the virus is transmitted and is a common factor for all entries of *ℬ*, (ii) an age specific susceptibility factor *σ*^*A*^ that describes the average biological susceptibility to the virus and the average activity level (in terms of social contacts) of the given age group, *A*, and is a common factor for each row of *ℬ*, (iii) the effective infectivity 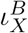 of each given compartment *X* and age group *B* that depends on the activity level and the plain infectivity of *X*^*B*^, and is a common factor in each column of *ℬ*, and finally (iv) a likeness factor *ϕ*^*BA*^ that describes the mixing of age group *B* with age group *A*.

The factor *β*_0_ has to be estimated by fitting the model to data provided by the RKI [31]. The same is true for the age specific susceptibilities that depend on the immune response of the individual (supposedly on average stronger in younger individuals).

The effective infectivity is supposed to be lowest for diagnosed individuals (*H*) thanks to deliberate contact reduction. We also assume symptomatic individuals to be less active than asymptomatic ones which means that 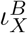 is smaller for *X* = *I* than for *X* = *U* even though the biological infectivity should be higher for symptomatic individuals than for asymptomatic ones. This activity reduction results from both the symptoms restricting the mobility of an affected individual as well as them being aware of their potential to spread a communicable disease. We also assume individuals in the late exposed stage *E*_3_ to be highly infectious which is in keeping with studies on the transmission dynamics of SARS-CoV-2 as exemplified by [17].

The likeness factor is hard to estimate. Following [26] who have done so for the population in Hubei province in China, we therefore consider contacts in three different realms: at home, at work and school, and in other locations, most notably during leisure activities. For each activity, we assume specific contact distributions depending on the ages of the individuals. For example, we assume high contact rates among adults and between adults and children at home. The separation of the realms allows for each intervention to produce a specific effect. For example, school closures predominantly act on the child-child contacts in the school realm. The resulting matrix *B* under the influence of different interventions is graphically illustrated in Fig. 4.

The progression rates *γ*_*X*_ are determined by the estimated mean durations of each stage of the infection. For simplicity, and analogously to [5], we choose all the rates for a given compartment to be the same (that is, *γ*_*E*1_ = *γ*_*E*2_ = *γ*_*E*3_ =: *γ*_*E*_ and likewise for *U, I*, and *H*). In accordance with [23] we assume the mean incubation period to be 5.5 days and therefore put *γ*_*E*_ = 3*/*5.5, the factor 3 giving a mean combined duration of stay in *E*_1_, *E*_2_, and *E*_3_ of 5.5 days. In the same manner we take the rates *γ*_*U*_ = 3*/*7 and *γ*_*H*_ = *γ*_*I*_ = 3*/*8 corresponding to mean durations of the infectious period of seven days for asymptomatic individuals and eight days for symptomatic ones, in accordance with previous studies [7, 26].

The probability *ρ*_0_ of developing symptoms at the end of the incubation period is estimated to be 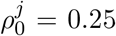 for juniors who are reportedly often asymptomatic, and 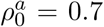 and 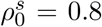 for adults and seniors, respectively.

The probability of an infection being discovered by the end of incubation the period is called *η*_0_ and is assumed to be small in the beginning of the simulation. The same is true for the rates 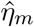 for discovering an infection in the absence of symptoms. For symptomatic individuals, these rates are larger as symptoms provide a strong suspicion of being infected. We assume a certain age dependence for testing. On the one hand, seniors are at high risk and might be expected to be tested more frequently but on the other hand, severe respiratory tract infections are common in people with limited immune competence and may not be taken as implicating a SARS-CoV-2 infection. Juniors are reportedly not seriously affected, and we assume that, therefore, they are less frequently tested.

According to numbers provided by the RKI [32], testing has been dramatically increased between March 9 and March 15, so we assume that all the testing rates mirror this increase. In particular, we assume the rates *η*_0_ and 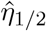 to be significantly larger than zero since individuals are supposed to be tested even in in the absence of symptoms if they were in close contact with a known infective.

The parameters *ν*_*m*_ describe the probability of progressing to the next stage while being diagnosed (i.e. transitioning from an *I* to a *H* compartment). They are assumed to be close to 1, them being smaller would imply the assumption that being tested is somehow associated with more severe symptoms and would, therefore, slow down the progression toward recovery.

To follow weekly oscillations in the reported cases, which show a regular slump over the weekends, we include time dependent test rates. This allows the simulated case numbers to closely follow the data as illustrated in Fig. 6b. For long term simulations of scenarios we assumed these fluctuations to fade away and removed the time-dependency from the testing rates. This implicitly represents an assumed increase in testing activities, and de facto increases the reporting ratio by more than 10%.

### Data

The publicly available dataset provided by the Robert Koch Institute (RKI) [31] was used for this study. Statistical analysis of the dataset was performed (results not shown here) analogously to our previous work [4].

## Footnotes

1 The leading relevant eigenvalue of the linearization about the equilibrium with only susceptible individuals is located close to the imaginary axis.

